# S-variant SARS-CoV-2 is associated with significantly higher viral loads in samples tested by ThermoFisher TaqPath RT-QPCR

**DOI:** 10.1101/2020.12.24.20248834

**Authors:** Michael Kidd, Alex Richter, Angus Best, Jeremy Mirza, Benita Percival, Megan Mayhew, Oliver Megram, Fiona Ashford, Thomas White, Emma Moles-Garcia, Liam Crawford, Andrew Bosworth, Tim Plant, Alan McNally

## Abstract

Birmingham University Turnkey laboratory is part of the Lighthouse network responsible for testing clinical samples under the UK government ‘*Test & Trace’* scheme. Samples are analysed for the presence of SARS-CoV-2 in respiratory samples using the Thermofisher TaqPath RT-QPCR test, which is designed to co-amplify sections of three SARS-CoV-2 viral genes.

Since more recent information became available regarding the presence of SARS-CoV-2 variants of concern (S-VoC), which can show a suboptimal profile in RT-QPCR tests such as the ThermoFisher TaqPath used at the majority of Lighthouse laboratories, we analysed recently published data for trends and significance of the S-gene ‘dropout’ variant.

Results showed that:

i. the population of S-gene dropout samples had significantly lower median Ct values of ORF and N-gene targets compared to samples where S-gene was detected
ii. on a population basis, S-gene dropout samples clustered around very low Ct values for ORF and N targets
iii. linked Ct values for individual samples showed that a low Ct for ORF and N were clearly associated with an S-dropout characteristic
iv. when conservatively inferring relative viral load from Ct values, approximately 35% of S-dropout samples had high viral loads between 10 and 10,000-fold greater than 1 × 10^6^, compared to 10% of S-positive samples.

This analysis suggests that patients whose samples exhibit the S-dropout profile in the TaqPath test are more likely to have high viral loads at the time of sampling. The relevance of this to epidemiological reports of fast spread of the SARS-CoV-2 in regions of the UK is discussed.

## Methods and data

The data used for this analysis are RT-QPCR threshold-crossing (Ct) values originating from Birmingham Turnkey Lighthouse lab testing under the UK Department of Health and Social Care *‘Test and Trace’* network. The laboratory installed at the University of Birmingham - the design and operation of which has been described earlier [1] relies on the Thermofisher TaqPath RT-QPCR test [2].

A dataset of positive results from the first months of operation was released as a supplementary table in a recent publication comparing results of lateral flow device testing with RT-QPCR [3]. Raw Ct values from the supplementary table were analysed with respect to the presence of ORF-, N-, or S-gene target undetectable signals. To assist the analysis, true target-negative results – defined as ‘no signal detected’ rather than a Ct value above the cut-off value of 37 - were given a nominal Ct value of ‘45’.

Frequency comparisons, Chi-squared, and Mann-Whitney ‘U’ tests for significance of non-Gaussian distributions between S-gene negative and S-gene positive RT-QPCR results were performed using GraphPad Prism version 5.03.

## Results

### Frequency analysis of TaqPath Ct values from SARS-CoV-2 positive samples

Figure 1 shows the distribution of Ct values for the viral ORF gene across the range of positive samples, which revealed a possible multiphasic distribution of sample results for ORF and N gene detection (latter not shown). This was most obvious with a nadir in Ct frequency between 22 – 24.

**Figure 1:**
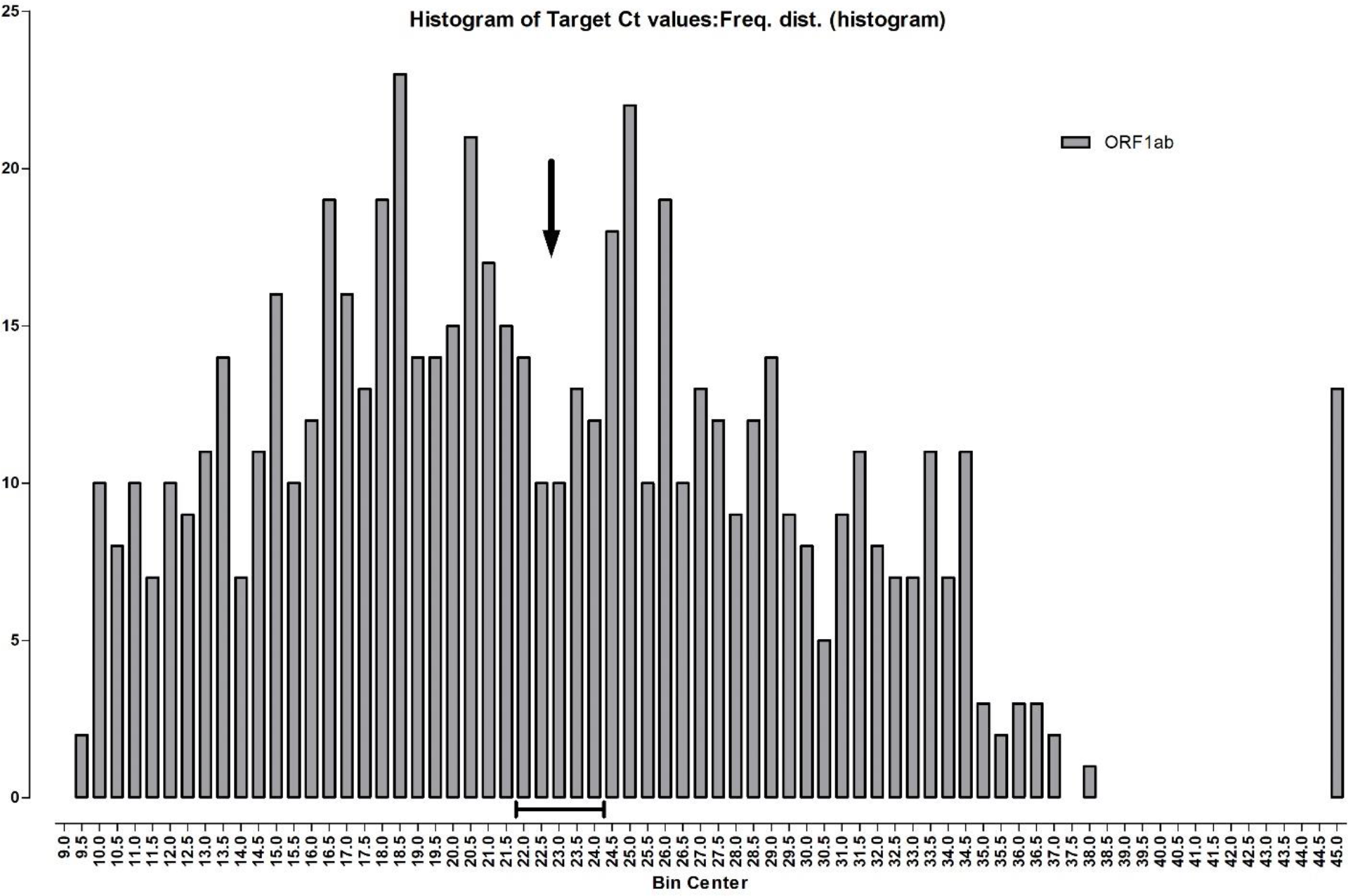
Simple frequency analysis of ORF gene Ct values from 641 positive samples, with a bin size of 0.5. Arrow and bar on the X-axis indicate nadir of frequency and Ct values, respectively. A Ct of 45 is displayed in the analysis and denotes “no signal detected” in the assay.

### Frequency analysis for all three gene targets

Figure 2 showed a high proportion of S-gene negative samples; note the number having an S-gene undetectable profile (178 of 641; 27.7%) at far right, compared with ORF and N-gene undetectable positive profiles (both 13 of 641; 2.0%).

**Figure 2:**
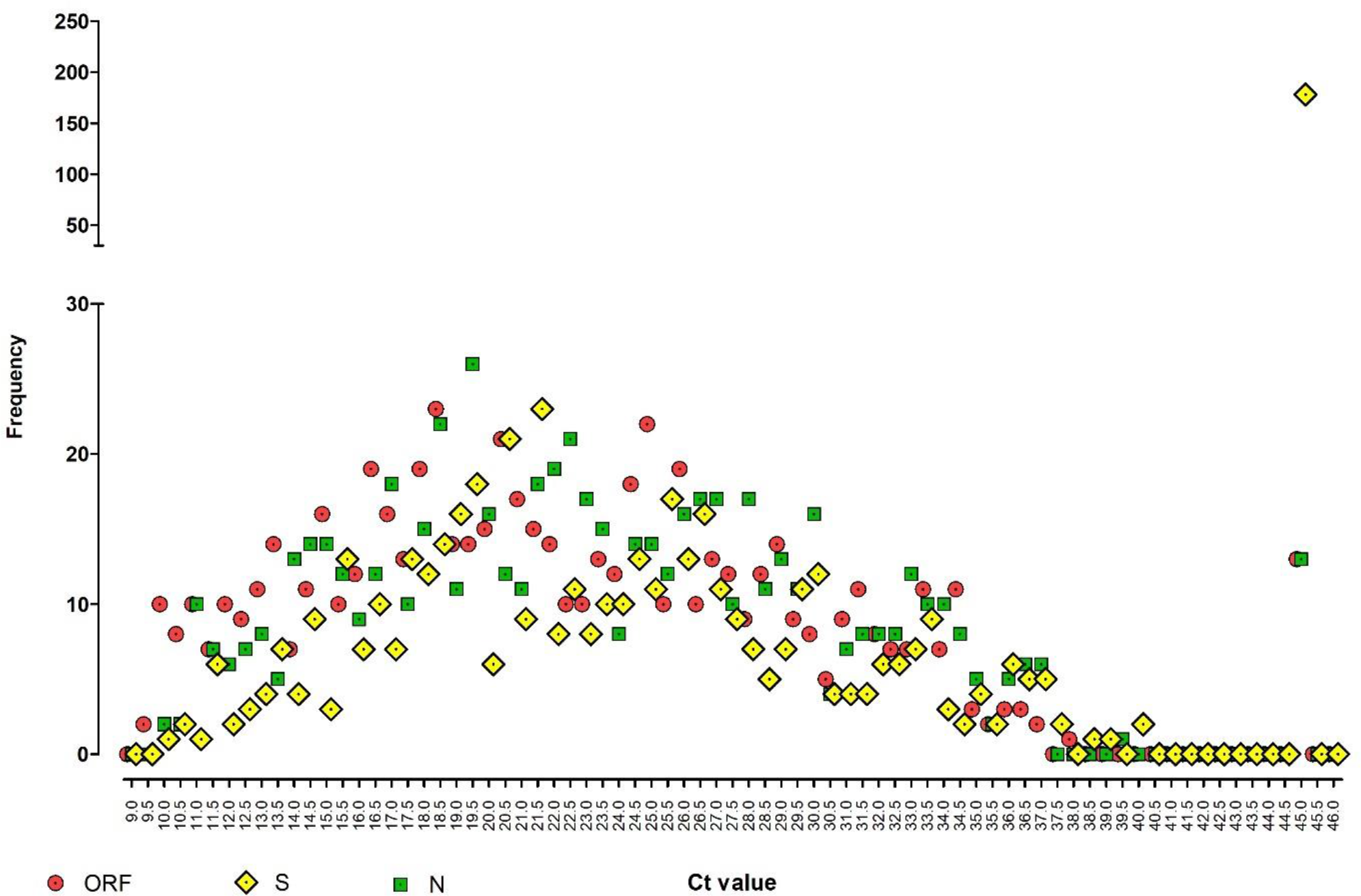
Dot-plot frequency analysis showing the relationship between three viral gene targets detected by the TaqPath RT-QPCR test, across the full range of Ct values. Y-axis is divided into a low frequency range of 0-30 and a high frequency range of 50-250. A Ct of 45 is artificially assigned to represent results where no signal was detected in that assay.

Further observation of the profile in figure 2 shows that lower Cts (9 – 22) for ORF and N tend to have a lower frequency for the corresponding S-gene, but at CTs from 25 -33 and onwards, this trend is less obvious with similar frequencies of all three viral genes detected. In particular, at the extreme end of the test sensitivity range (Ct 33 – 40), ORF, N, and S are detected at approximately equal frequencies with no apparent loss of sensitivity for any gene target.

### S-gene undetectable samples had significantly lower median Ct than ORF- and N-gene

To further determine whether the observation that S-gene undetectable samples were significantly associated with lower Ct values of ORF and N gene positive samples, the distribution of S-negative and S-positive samples was compared within all ORF and N positives (figure 3). In both, the median Ct value of S-negative and S-positives was significantly different; see table 1 (both p < .0001).

**Table 1:** Sample numbers and median (plus upper/lower quartiles) of positive sample Ct values for TaqPath RT-QPCR. Results are segregated for S-gene negative or positive samples, for each group where either the ORF or N-gene was positive. In both ORF and N-gene groups, the median Ct of S-negative and S-positive targets was significantly lower (Mann-Whitney ‘U’; two-tailed; p < .0001). In each case, the difference in median Ct represents a 10 - 100-fold higher viral load in the S-negative group.

|  | ORF-pos |  | N-pos |  |
| --- | --- | --- | --- | --- |
|  | S-neg | S-pos | S-neg | S-pos |
| Number of values | 178 | 450 | 178 | 450 |
| Minimum | 9.47 | 9.99 | 10.02 | 10.53 |
| 25% Percentile | 13.15 | 18.24 | 14.28 | 19.30 |
| Median | 18.16 | 22.30 | 19.39 | 23.16 |
| 75% Percentile | 26.74 | 27.25 | 28.08 | 28.12 |
| Maximum | 36.49 | 37.94 | 36.73 | 39.31 |

**Figure 3.**
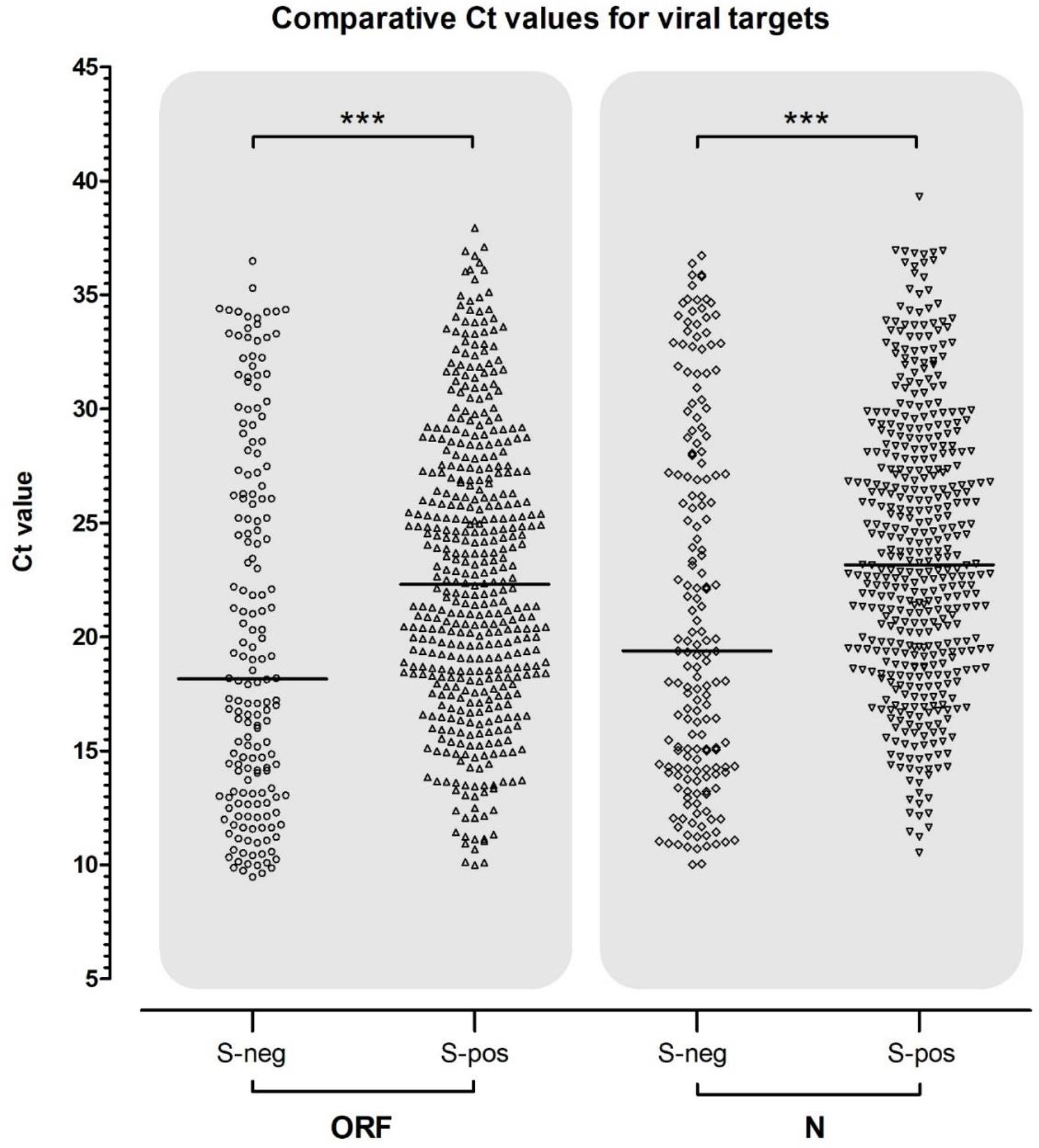
Scatter plot of the population of S-gene negative and positive Cts within corresponding ORF and N-gene positive samples. Median Ct is shown by a black horizontal bar and above both plots the results of tests for significant differences are shown with conventional notation.

**Figure 4:**
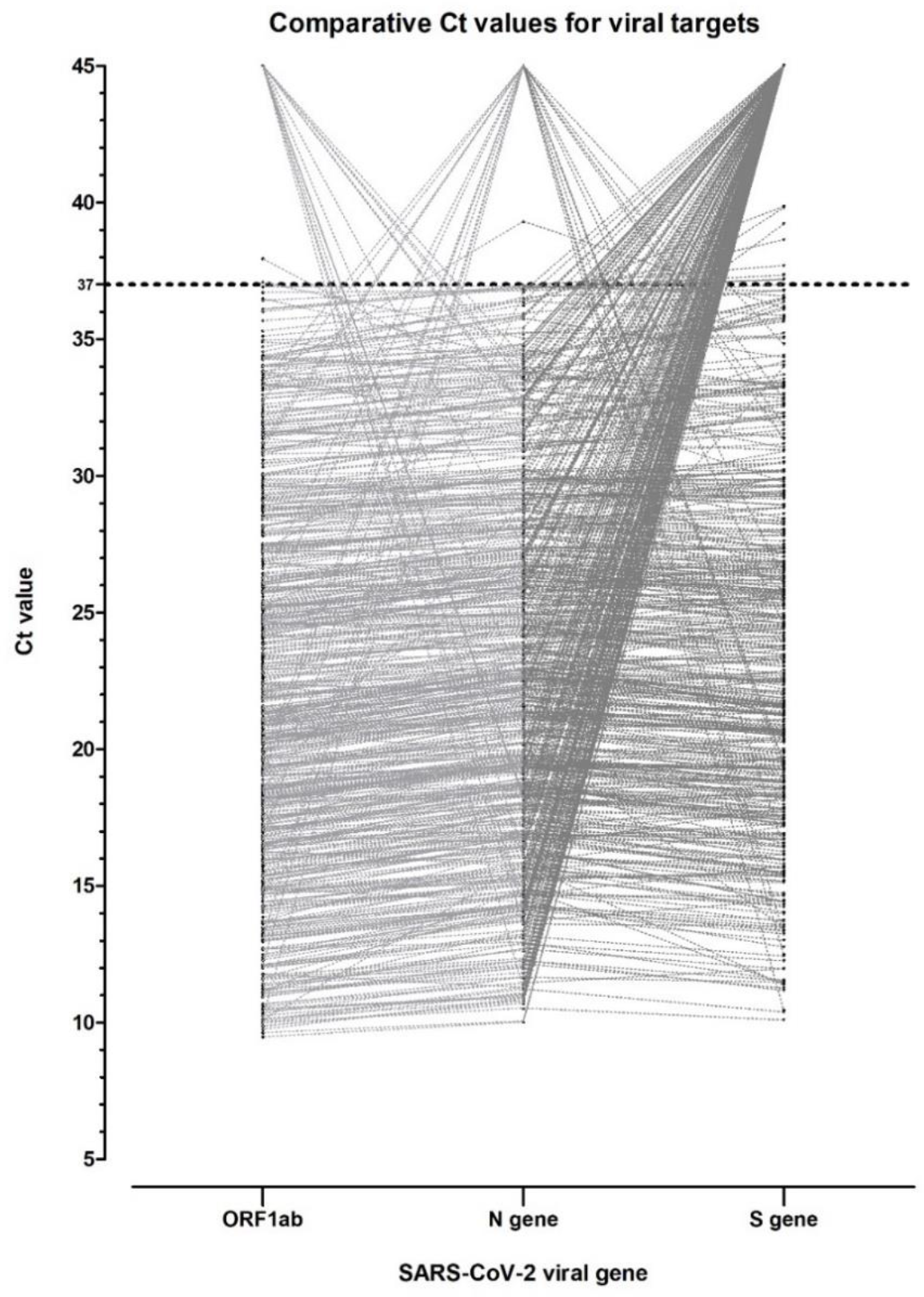
Scatter plot of viral gene Ct values for individual positive samples. Each set of 3 Ct values for a sample is linked by a grey dotted line. A Ct of 45 is shown in this analysis which represents test where no signal was detectable in that test.

Although S-negative samples occurred across the range of Ct values for ORF and N genes, clustering of S-negative results around very low Ct values of ORF and N can be clearly seen, and probably accounts for the lowering of the median Ct

### Comparisons of linked Ct values for individual samples show a propensity for S-negative samples to be associated with a lower Ct for ORF and N gene targets

We investigated the relationship within each positive sample for the S-gene negative profile to be associated with low Ct values of ORF and N gene targets. Although S-negative samples did occur across the range of Ct values, they appear most pronounced towards the lower end of the range. In case this observation was a graphical artefact, we performed a Chi-squared analysis of the number of samples below a Ct value of 15 in both S-negative and S-positive categories in the ORF-positive group. This analysis showed a highly significant difference between proportion (*X*^2^ (1, *N* = 628) = 36.61, *p* < .001).

Some of the 13 ORF and N-gene negative samples were also associated with low Ct values, and should be further investigated for possible mutations interfering with specific amplification of the target.

## Discussion

A major part of the UK response to the spread of SARS-CoV-2 through the population has been the setup and operation of several UK centres performing high-throughput RT-QPCR testing. These have been standardised by adopting an RT-QPCR test process around the ThermoFisher ‘TaqPath’ test. The test sensitivity and specificity are verified between centres using the same Qnostics external quality assurance panel [4] and internal quality control maintained through standard techniques such as Levey-Jennings and Westgard criteria.

Our analyses of data, generated during the first months of Birmingham Lighthouse Turnkey operation, shows a high proportion of SARS-CoV-2 positive samples which had the viral S-gene target undetectable with two other gene targets ORF and N clearly detected (termed ‘S-dropout’ samples). Further analysis shows that a significant proportion of S-dropout samples are associated with lower Ct values of ORF and N in the same sample; from which it possible to infer a relatively higher viral load in these specimens.

Clearly, the higher viral loads inferred from S-dropout samples could determine the infectiousness of subjects, and thus the ability of the virus to transmit onwards. The significant difference in population median Ct value, between S-dropout and S-detected samples, represents between 10 and 100-fold increase in target concentration for S-dropout. The cluster of S-dropout samples having ORF and N Ct of between 9 and 15 (63/178 (35.4%); 46/450 (10.2%), respectively) is a corresponding further increase in relative viral load of between 10 and 1,000-fold.

Based on our Qnostics EQA verification data for the TaqPath test (table 2), a Ct value of approximately 15-16 corresponds to a viral load of 1 × 10^6^ copies per millilitre (mL). Therefore, our observed cluster of S-dropout samples at Ct less than 15 corresponds to a conservative estimate of a significantly larger population of infectious subjects that have an increased viral load up to 10,000-fold higher. Such capability of increased transmission has been ascribed to an S ‘variant of concern’ apparently spreading throughout the South-east of the UK, and possibly beyond; but epidemiologically it is difficult to disentangle other reasons for more efficient spread, such as human behavioural factors, from those of a clinical virological nature.

**Table 2:** Verification data from the Lighthouse Turnkey laboratory, relating copies per mL of inactivated SARS-CoV-2 viral lysate in the EQA sample dilution series, to Ct values obtained for different viral gene targets amplified in the TaqPath test.

| QNostic Sample ID | Copies/ml | Log<sub>10</sub><br>c/mL | Mean C<sub>t</sub> of 6 replicates (% agreement) |  |  |
| --- | --- | --- | --- | --- | --- |
|  |  |  | Orf1ab | S Gene | N Gene |
| SCV2AQP01-S01 | 1000000 | 6 | 15.3 (100%) | 16.5 (100%) | 15.8 (100%) |
| SCV2AQP01-S02 | 100000 | 5 | 18.3 (100%) | 20.5 (100%) | 17.4 (100%) |
| SCV2AQP01-S03 | 10000 | 4 | 22.8 (100%) | 24.9 (100%) | 23.6 (100%) |
| SCV2AQP01-S04 | 5000 | 3.7 | 19.5 (100%) | 24.8 (100%) | 24.1 (100%) |
| SCV2AQP01-S05 | 1000 | 3 | 25.9 (100%) | 28.6 (100%) | 26.6 (100%) |
| SCV2AQP01-S06 | 500 | 2.7 | 25.8 (100%) | 29.1 (100%) | 25.8 (100%) |
| SCV2AQP01-S07 | 100 | 2 | 27.7 (33%) | 30.3 (66%) | 30.8 (100%) |
| SCV2AQP01-S08 | 50 | 1.7 | 29.3 (17%) | 31.1 (27%) | 29.1 (55%) |
| SCV2AQP01-S09 | Negative | ---- | Not detected | Not detected | Not detected |

We considered alternative explanations for S-dropout positive results at very low ORF and N Ct values. One is that chemical components in an individual RT-QPCR reaction run short when amplifying multiple targets at high viral load input, and possibly the S-gene target is first to become non-amplifiable. However, the TaqPath test contains an internal control provided by co-amplification of non-human bacteriophage MS-2, the target RNA of which is included at a concentration that is more likely to become undetectable under adverse reaction conditions than any of the three specific gene targets; and which provides reassurance that amplification of all specific targets is not being inhibited. All positive TaqPath results in the data table were passed as valid, as determined by the presence of MS-2 amplification, making the likelihood of S-gene dropouts being due to a general reaction chemistry bias as highly unlikely.

Secondly, during the laboratory verification stage of the TaqPath test using a nationally accepted EQA standard dilution series which consists of cultured, inactivated whole SARS-CoV-2 RNA, there was no observed behaviour of the S-gene target to drop out at Ct levels of approximately 15; which is lower than the median Ct value at which S-gene dropouts were seen to occur in our data. This prior observation during verification is a second reason to discount the influence of general RT-QPCR reaction inefficiencies in our observations.

It should be emphasised in this report that the authenticity of results for individual samples is not affected by the presence of the S-dropout phenomenon, as the TaqPath RT-QPCR test result is classed as ‘positive’ when at least two gene targets are detected. Thus, the ability of the TaqPath test to detect three viral targets provides a degree of robustness to the ‘*Test and Trace’* programme, even when a viral mutation renders one of them undetectable. The test may provide a useful means to track the spread of the S-VoC and monitor for the appearance of other gene mutations in the population which may affect the accuracy of other diagnostic tests. We observed clear dropouts for the other ORF and N genes – albeit at a much lower frequency and not apparently associated with high viral loads – and we believe these should be similarly investigated for mutations in the corresponding genes that could have affected their detection in the TaqPath test. We also note that double-dropouts – where two viral genes are not amplified in a sample –by implication will not be represented here as they would be classed as a negative result and not included in the dataset. A more exhaustive analysis would involve reviewing all negative results with a single viral gene amplified.

Limitations of this data are:

a. Our analysis provides supporting evidence to explain why the S-VoC may be transmitting more rapidly amongst populations, but it does not provide an explanation of how an increased viral load could occur. The biological plausibility of higher infectivity, whether through evolutionary viral replication advantages or evasion of the host immune system, will become apparent through infectivity studies of currently being performed.
b. Although we have made broad inferences in relative viral load in the samples, the TaqPath is not a quantitative assay for SARS-CoV-2 and our observations should be repeated by a dilution series or a validated quantitative method.
c. Our investigation does not directly link the RT-QPCR S-dropout phenomenon to specific viral mutations known to interfere with RT-QPCR detection, and whole genome sequencing of individual samples will prove valuable in strengthening the association with changes in the viral genome.

At the time of writing, temporal or geographic data is not available for individual samples included in this analysis. Clearly, other Lighthouse centres, NHS or private sector laboratories using the TaqPath test, or indeed other RT-qPCR involving S-gene detection, should add data to this finding to confirm or refute the analysis performed here.

## Data Availability

Data used in this analysis has been previously published as a supplementary file, and is referenced as such in the manuscript

